# NSTEMI Decide: Development of a Decision Aid for Older Adults with Non ST Elevation Myocardial Infarction

**DOI:** 10.1101/2021.07.29.21261343

**Authors:** Jenny Summapund, Rachel A. Sibley, Sohah Iqbal, Nicholas J. Kiefer, Aisha Langford, Erica Spatz, Mallory Barnett, Joshua Rivers, Sarwat I. Chaudhry, Victoria V. Dickson, Nikhil V. Sikand, Daniel D. Matlock, John A. Dodson

## Abstract

**Background:** Many patients hospitalized with non-ST elevation myocardial infarction (NSTEMI) are older adults (age ≥75), in whom invasive coronary angiography confers potential benefits but also higher risks than the general population. Our goal was to therefore develop a patient decision aid (PtDA) to assist with shared decision making (SDM) between cardiologists and older adults with this condition.

**Methods:** We followed International Patient Decision Aid Standards to develop the *NSTEMI Decide* PtDA. The initial prototype was based on structured interviews from 20 patients and 20 clinicians. Risks and benefits of invasive coronary angiography were derived from the available literature in older adults, with an emphasis on findings from randomized trials. The PtDA was then revised through an iterative, user-centered design approach with rapid prototyping using input from both clinician and patients to meet the needs of both stakeholders.

**Results:** The PtDA went through 8 iterations. The final decision aid was 11 pages and included background information (6 pages), explanation of benefits and risks (4 pages), and a summary (1 page). Large font (≥26 point) was used to accommodate visual difficulties. Language was tailored to an 8^th^ grade reading level. Based on the best available literature, we included benefits of invasive coronary angiography (recurrent myocardial infarction, repeat revascularization) and risks (stroke, bleeding, acute kidney injury). The final PtDA was converted to a mobile health application to increase usability on portable digital devices in the clinical setting.

**Conclusions:** We developed the first PtDA tailored to older adults hospitalized with NSTEMI. This PtDA will be prospectively tested to evaluate dimensions including feasibility of use, satisfaction with care, medical knowledge, and decisional conflict.

## INTRODUCTION

Shared decision making (SDM), which involves the active participation of patients in health care decisions that have multiple acceptable choices,^1^ is a key part of achieving a more patient-centered health system. The basic tenet of SDM is that clinicians are experts in medical evidence, while patients are experts in what matters most to them.^2^ In cardiovascular medicine, where invasive procedures are common, SDM has been recognized by professional societies^3^ and payors^4^ as an integral component of a more patient-centered health system.

Patient decision aids (PtDAs) are a tool that can be used at the point of care to encourage SDM by facilitating conversation, empowering patients, and providing a more thorough review of treatment options outside the routine clinical encounter.^5^ Randomized controlled trials across many settings have shown that these tools increase patients’ knowledge (including understanding of risk), increase concordance between patient values and decision choice, decrease patients’ decisional conflict,^6,7^ decrease patients’ anxiety and depression,^8^ and lead to more accurate expectations about treatment course and possible complications.^2,7^ To date, in cardiovascular medicine, PtDAs have been developed for implantation of left ventricular assist devices,^9^ implantable cardioverter defibrillators,^10^ and left atrial appendage occlusion devices.^11^ Among older adults, SDM is especially relevant as surveys repeatedly demonstrate that they often prioritize health outcomes other than simply prolonging life,^12,13^ and treatment-related harms weigh heavily on their willingness to accept therapies.^14^

While non-ST elevation myocardial infarction (NSTEMI) requires major management decisions – most importantly, whether to pursue invasive coronary angiography – to date a PtDA has not been developed. Decision making around invasive coronary angiography is especially nuanced in older adults who have the potential to benefit from this procedure but are also at greater risk of harm than younger patients. For example, in a trial of invasive coronary angiography versus medical therapy for NSTEMI, patients age ≥75 were over three times as likely to experience major bleeding with invasive coronary angiography.^15^ Notably, most trials of invasive coronary angiography for NSTEMI were in younger patients or older adults thoroughly screened to exclude major comorbidities: for example, in a frequently cited meta-analysis of 7 studies evaluating the benefit of a routine invasive strategy in patients with NSTEMI, the mean age was 62 years.^16^

In the context of previous described benefits of PtDAs and clinical uncertainty in managing older adults with NSTEMI, we developed the first PtDA for older adults with NSTEMI considering invasive coronary angiography. This manuscript describes the process for development of this decision aid, which we have entitled *NSTEMI Decide*.

## METHODS

### Overview

The Ottawa Decision Support Framework proposes that decision support should be a direct response to a person’s decisional needs thus improving the quality of decisions and ultimately their behavioral and health outcomes.^17^ The development process for *NSTEMI Decide* followed the International Patient Decision Aid Standards (IPDAS) Collaboration criteria,^18^ an evidence-informed expert consensus guidance regarding the aspects of decision aids that lead to better decisions.

A multidisciplinary team was created to steer development of the PtDA. The development team consisted of an interventional cardiologist (SI), a noninvasive cardiologist (JAD), a geriatrician (DDM), a nurse practitioner (VVD), an internist (SIC), and a research coordinator (JS). All aspects of the study were approved by the NYU Grossman School of Medicine Institutional Review Board, and either verbal or written informed consent was obtained from participants prior to study involvement.

### Needs Assessment Among Patients and Physicians

Guided by the principles of the Ottawa Framework, our first step in PtDA development was conducting a needs assessment among patients and clinicians to understand their perspectives on SDM and identify current gaps in their decision-making process, including patients’ decisional conflicts (uncertainty in knowledge, support, or confidence that would ultimately lead to decision regret),^19^ as well as clinicians’ perspectives on real-world practice.^20^ For clinicians, we specifically targeted cardiologists since in practice they are most directly involved in decision making regarding invasive coronary angiography.

To evaluate decision needs among patients, two study coordinators conducted one-on-one qualitative telephone interviews with 20 patients (age ≥70) who had recently experienced an acute myocardial infarction (AMI) at NYU Langone Health (NYULH). Questions focused on identifying the factors underlying patients’ decisions to undergo a coronary revascularization, prior knowledge or experience of the procedure, and decisional needs. To elicit clinicians’ views on SDM support needs, the study coordinators performed similar in-depth qualitative interviews with 20 cardiologists employed by NYULH with focus on exploring their treatment strategies among older adults with NSTEMI, how they defined the term “shared decision making,” and their needs for a tool to facilitate SDM discussions in a clinical setting. Interviews were transcribed verbatim, coded using Atlas.ti software, and analyzed using a ground-theory approach. Findings from these interviews have been previously published^21^ and were used as foundational work for the current PtDA development.

### Synthesis of Evidence - Outcomes

We conducted a review of the literature to synthesize the current evidence base of treatment options available for older adult patients presenting to the hospital with NSTEMI. Studies were selected that compared outcomes between an invasive approach (invasive coronary angiography) and a conservative approach for this condition. We searched two databases (PubMed and Google Scholar) using different combinations of the terms: “NSTEMI,” “treatment,” “coronary revascularization,” “elderly”, “geriatric,” “outcomes,” and “complications,” with a limit applied to studies published within the prior ten years. While limited by the relative paucity of high quality trials conducted specifically in older adult populations,^22^ our review yielded a total of 4 randomized controlled trials, 1 meta-analysis, and 10 retrospective cohort studies. Findings from these studies are summarized in Supplemental Table S1.

### Decision Aid Design

Per IPDAS guidelines, additional literature search was conducted by a research coordinator (JS) on health literacy, plain language usage, design, and graphic display.^23–26^ Design of past medical education materials that incorporate the use of plain language at the CDC-recommended 8^th^ grade reading level have been shown to be successful in both readability and usability among patients, especially older adults with lower health literacy.^27^

The resulting medical and design literature were then presented and thoroughly discussed among the core development team during regular meetings. Discussions focused on available treatments and potential complications, risk estimates, design, language, and feasibility in real-world practice. Discussions were documented, and these concepts were ultimately incorporated into the initial prototype development.

### Initial Prototype Development

Guided by the needs assessment, literature review, and IPDAS criteria, we developed the first prototype of the *NSTEMI Decide* PtDA for distribution and discussion among all stakeholders. We focused our developmental approach on 3 major attributes previously outlined by Matlock et al.^20^: (1) correct and accurate content, (2) readability and usability, and (3) acceptability and lack of bias. The initial prototype comprised 5 sections: an illustrated overview of the medical condition (AMI), a module of the two broad treatment options (invasive vs. conservative management), a clinician-facing checklist to facilitate SDM discussions, a discussion page addressing patients’ emotions and values; and a comparative module presenting the benefits and risks of each treatment option. To facilitate ease in editing, we determined that a paper-based PtDA format was most appropriate during this initial prototype phase. Paper-based PtDAs have shown to be preferred by patients over more interactive decision tools in selected settings.^28^

The first prototype was presented at our standing developmental team meeting. Modifications were made primarily to the medical content, including refining the accuracy of risk estimates and amending illustrations for improved clarity. Edits to the design were also made in order to follow user-design principles,^29^ with concentration on feasibility of application in a clinical setting for clinician users, and readability for patient users.

### Successive Iterations to PtDA

Successive drafts with modifications to content and design of the PtDA were generated based on feedback from the initial prototype. To continuously assess the acceptability, usability, and preliminary usefulness of our PtDA, we serially presented our iterations to patient and clinician advisory groups as described below.

#### Patient Advisory Group (General)

A fifth version of our PtDA was presented to the NYU Patient Advisory Council for Research (PACR) to ensure readability and comprehension. PACR consisted of NYU patients whose eligibility for membership was facilitated by the NYU Clinical and Translational Science Institute for purposes of inclusivity and diversity. To ensure an equitable selection process, patients with at least 1 active medical condition were identified and recruited via electronic health record, and upon consent, were asked to participate in at least 6 meetings per year to review research projects.^30^ At the time of our focus group, PACR had 25 total members. Participants ranged from ages 18-78, with most in older age strata. Demographic information of PACR members are provided in Table 1.

**Table 1.** Patient Advisory Council for Research (PACR) Demographics.

|  |  |
| --- | --- |
|  | **N=25**<br>**(as of January**<br>**2019)** |
| **Sex** |  |
| Female | 14 |
| Male | 11 |
| ***Race/Ethnicity*** |  |
| Asian | 2 |
| Black | 12 |
| Hispanic | 3 |
| White | 8 |
| ***Age group, years*** |  |
| 18-36 | 6 |
| 37-55 | 3 |
| 56-74 | 12 |
| 75+ | 4 |

The focus group met for 1 hour and was moderated by a facilitator outside of the development team. Patients were asked to review the PtDA and comment on the overall content, design, language, graphics, and balance. Special emphasis was placed on the risk and benefits presentation and the group’s opinions on their understanding and interpretation of the statistics. Following the PACR session, members of the study team reconvened during a standing development meeting to discuss patient feedback and implement subsequent changes in the next iteration of the PtDA.

#### Clinician Advisory Group

To assess the validity of content and usability in a clinical setting, we then presented an updated iteration to investigators at the Colorado Cardiovascular Outcomes Research Consortium (CCOR). CCOR consists of clinician researchers with expertise in cardiology, geriatrics, emergency medicine, and public health.^31^ An electronic copy of the PtDA was sent to the CCOR coordinator and printed as paper copies in-person at the meeting for review, while feedback was provided via video conferencing. Attention was placed on medical content (such as risk estimates) and length in order to evaluate the accuracy and applicability of the tool in a clinical setting.

#### Patient Advisory Group (Geriatric)

To accommodate feedback from our target end-user patient population, our last focus group was conducted with a geriatric PACR subgroup who were presented our near-final version of the PtDA. Patients already participating in PACR age ≥65 were asked to participate in a second round of feedback, with particular focus on formatting design (such as font size and color), graphic illustrations, and comprehension of the risk estimates. Feedback on emotional impact and overall PtDA usability was also assessed among patients who had experienced a prior AMI.

Rounds of iterations and presentations between the clinician advisory group, patient advisory groups, and the development team were conducted until no new ideas emerged. Our final iteration of the PtDA was updated by the research coordinator (JS) with higher quality graphics and minor formatting modifications. For broader usability, two certified, native Spanish-speaking research coordinators translated the PtDA into Spanish.

## RESULTS

### CONTENT

The prototype went through 8 successive iterations over one year (1/2019-1/2020). The final *NSTEMI Decide* PtDA was 11 pages long; it consisted of 3 background pages, 1 page of a diagram depicting treatment options, 1 page addressing emotions and personal values, 4 pages of risks and benefits communication through pictograph and percentage display, and 1 summary page. Samples of the final PtDA are shown in Figure 1. The full version is available for download at https://aginghearts.org/more-information/geriatric-apps/.

**Figure 1.**
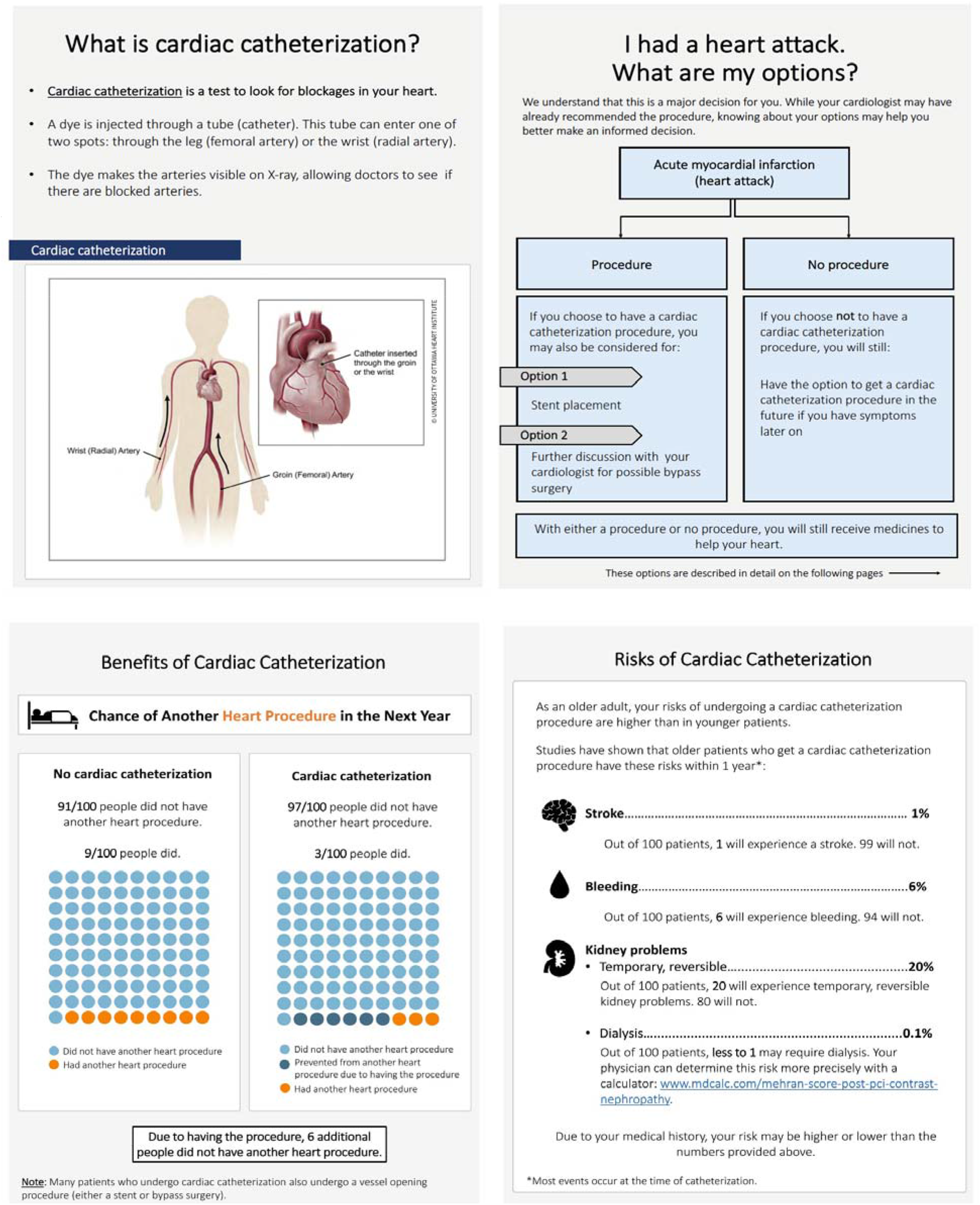
Sample Pages from NSTEMI Decide

#### Medical Background

A common concern among the patient advisory group was the nuances of medical terms used to describe the condition (AMI) and its related procedures. Concerns included the unknown differences between STEMI vs. NSTEMI, and interchangeable terms used to describe procedures, such as “percutaneous coronary intervention” (PCI) vs. “stent,” “cardiac catheterization” vs. “coronary angiography,” and “acute myocardial infarction” vs. “heart attack.” These terms, which are variably written in publicly available patient educational materials, contributed to the confusion patients reported when reviewing the material in our PtDA. To prevent overwhelming patients with medical terminology without compromising important information, we therefore provided 2 pages of background information filled with bolded terms, acronyms, and definitions; concise explanations of procedures; colored illustrations with clear labels (e.g. type of blood vessels, location); and consistency of terms used throughout the PtDA to ensure ease of use.

#### Personal Values and Emotions

In line with other PtDAs that provide patients with a “values clarification” opportunity and thus resulting in a higher proportion of patients choosing an option consistent with their values,^6,7^ we dedicated one page to acknowledging patients’ values and emotions. Emotions like fear and hopelessness are common among patients reaching a crossroads in decision-making during a perceived life-threatening situation, and may consequently play a major role in the decision-making process.^19,28^ Questions such as “How do I feel about procedures?” and “How do I see myself living out the rest of my life?” are multipurpose in acknowledging patients’ fear, inciting them to clarify their values, and initiating a discussion with their clinician.

#### Benefits/Risks

Driven by the needs assessment interviews indicating that a visual PtDA tool is best to convey tailored risks/benefits and increase choice awareness among NSTEMI patients,^21^ we designed our PtDA to visually present the benefits and risks of an early invasive strategy with cardiac catheterization as compared to an initial conservative strategy with medical management alone. We used data from clinical trials that included patients over age 75 to represent the target patient population intended for our PtDA (Table S1). To determine benefits of an early invasive strategy, we synthesized evidence from available randomized controlled trials and a meta-analysis, which consistently demonstrated a reduction in recurrent AMI and in repeat revascularization within one year of hospitalization.^32–35^ In terms of risks of an early invasive strategy, the three most common major complications identified in the literature were stroke, bleeding, and acute kidney injury. Due to a lack of consistent reporting in available randomized trials, retrospective studies were used to supplement our final estimates for these complications.^36–39^ These data were used to formulate final risk estimates. While synthesizing data from multiple sources to produce a final risk estimate for use in the PtDA is a complex process, final estimates were reviewed and validated based upon clinical experience by key stakeholders, including general and interventional cardiologists. Rather than using reference ranges, specific numeric risk estimates were used to quantify outcomes. Previous studies have shown that specific estimates (as opposed to ranges) are associated with more accurate risk perception and improved decision-making processes among patients.^18,20,40^

### DESIGN

Due to *NSTEMI Decide*’s target audience being the older adult population, special attention was given to the design and formatting of the PtDA to ensure the accessibility and comprehension of the material in this age cohort. Design was tailored toward older adults with potential visual impairments, using large font and simple pictures when feasible. A Sans-Serif typeface was used for easy-reading, with its size kept at a minimum of 26. Considerations for color-blind individuals were reflected in our usage of exclusively vibrant, blue and orange symbols when color is needed to highlight risk estimates. Language was tailored to an 8^th^ grade reading level to ensure inclusivity using CDC’s Healthy Literacy and Plain Language Guidelines.^27,41^

Throughout this process, discussions with all three advisory groups (PACR, geriatric patient advisory group, and CCOR) indicated that a pictograph was the preferred way to visually display risk estimates of the two different treatment options. Both patient advisory groups indicated they preferred visuals to a “confusing numbers and decimals” presentation. Clinicians similarly agreed that pictographs provide better comprehensibility among patients when presenting risk estimates. And as evidenced in a prior study by Choi et al.^42^, older adults with low health literacy have better recall when presented with pictographs compared to text-based information.

While we used pictographs to display the benefits of cardiac catheterization, we displayed risks (stroke, bleeding, and acute kidney injury) as percentages alone. This was done for several reasons: first, the absolute risk was relatively low, making pictograph display more challenging; second, the theoretical risk of these complications without cardiac catheterization was zero; third, according to a consensus document from IPDAS,^26^ risk is equally transparent when communicated in percentages as compared to pictographs. To further tailor the risk presentation to older adults, we embedded a hyperlink within the PtDA to an existing acute kidney injury risk calculator, developed by Mehran and colleagues^43^, to better assist clinicians in assessing individualized risk scores for patients.

Other components, such as minimalistic designs; vibrant, color-blind friendly colors; and large font were also a common preference among patients and clinicians. Patient advisory groups collectively preferred a more minimalistic presentation, indicating that they want the benefits and risks presentation to “tell me what I should be looking at immediately.” Choices in language and medical terminology, however, had more conflicting opinions from both sides (illustrated in Figure 2). The entire iterative process is detailed in Figure 3.

**Figure 2.**
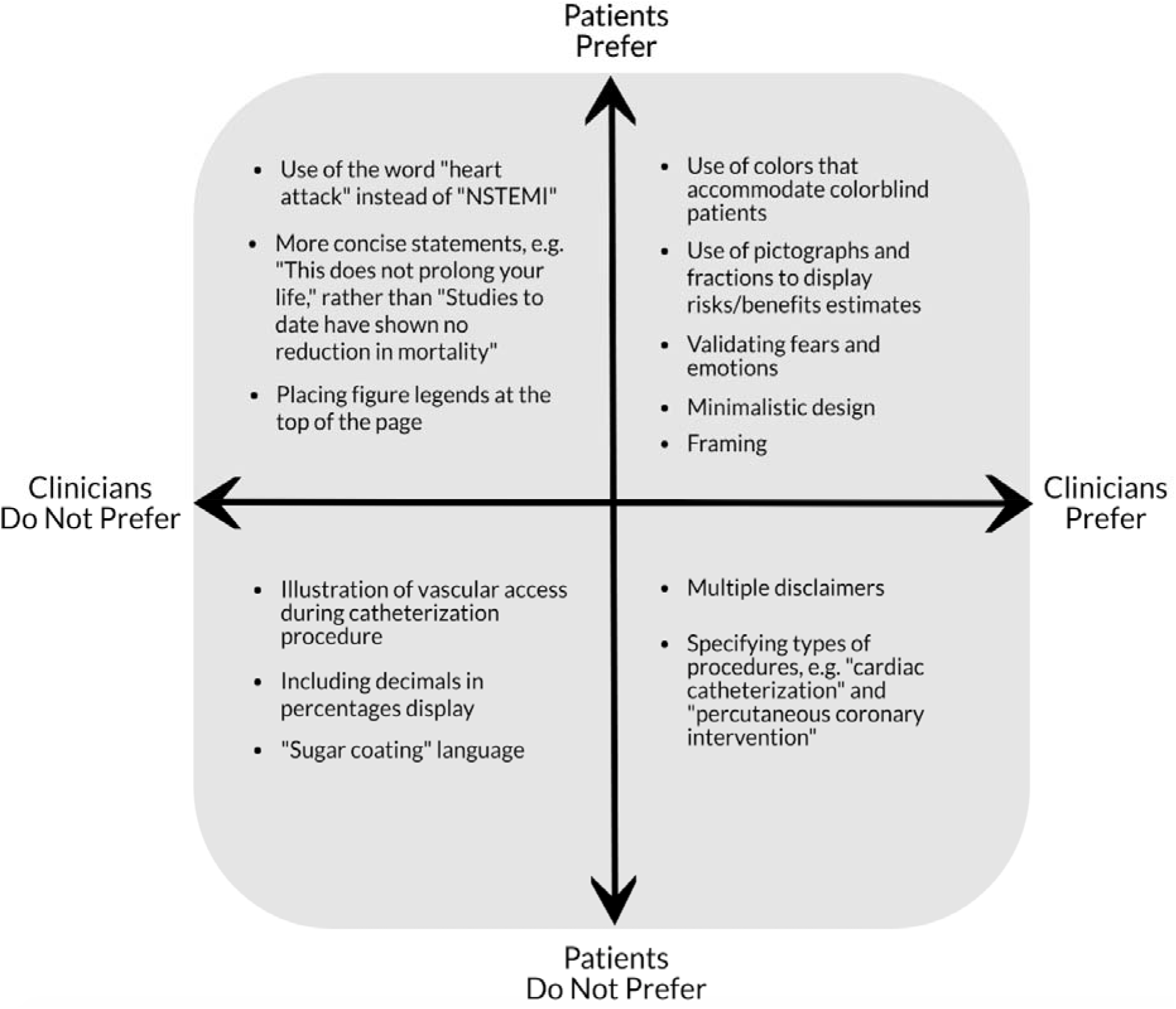
Sample Patient and Clinician Advisory Group Content and Design Preferences

**Figure 3.**
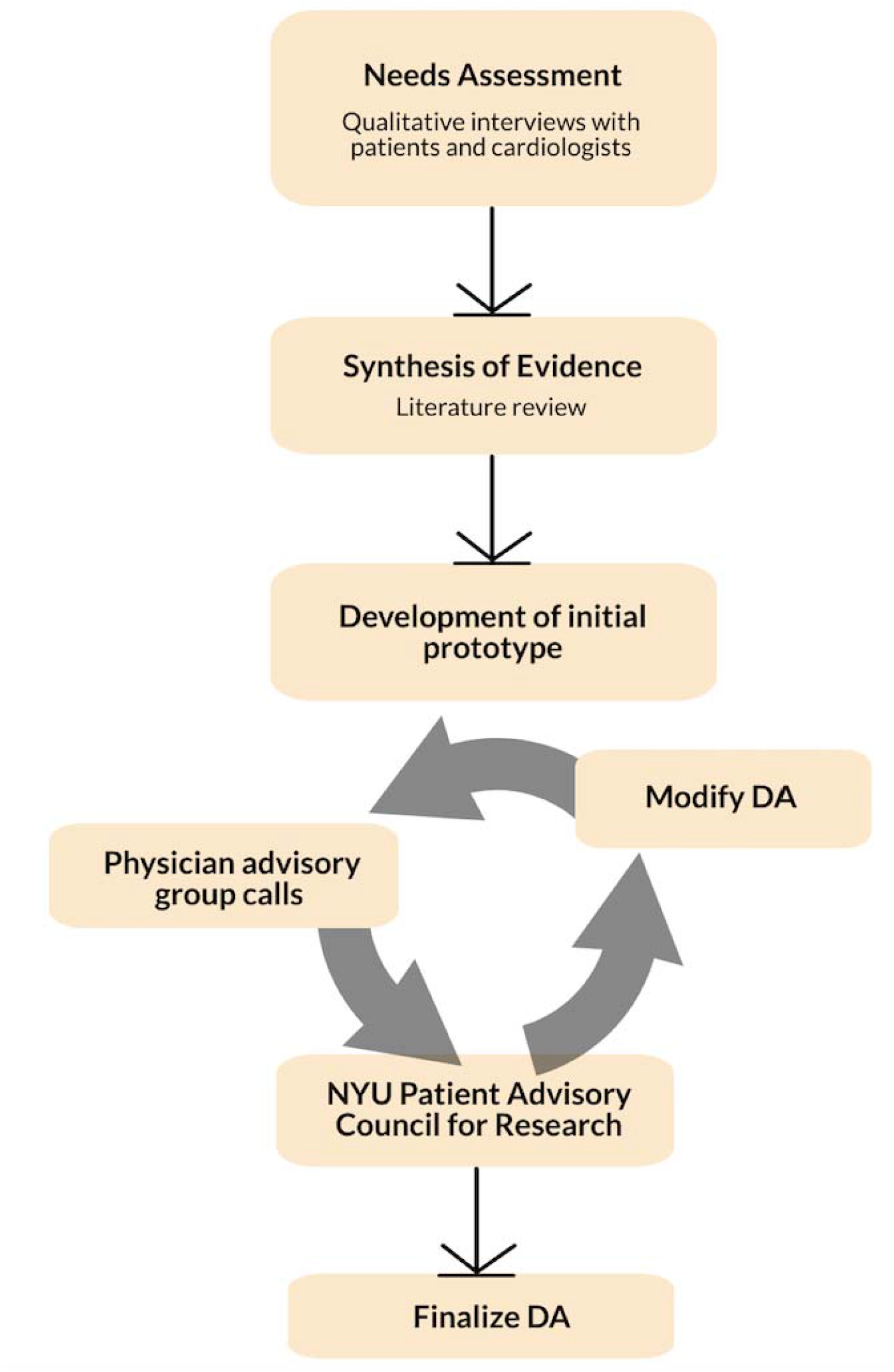
Development Process of NSTEMI Decide

### USABILITY

Physicians from the needs assessment interviews highlighted a desire for PtDA availability in the inpatient and bedside setting where clinical decisions for NSTEMI occur. Since all clinicians interviewed reported using portable electronic devices (smartphone and/or tablet) on a daily basis, and the logistics of printing a paper PtDA in the inpatient setting was deemed prohibitively time-consuming, we developed the final PtDA with digital implementation in mind. Accordingly, we had the final PtDA converted into an app usable in iOS format (an Android version is under development).

## DISCUSSION

As invasive coronary angiography becomes more common among older adults with NSTEMI, there is an imperative to provide appropriate information regarding its benefits and risks. Our needs assessment, based on interviews with both patients and cardiologists, demonstrated a need for tools to improve communication in the inpatient setting. We accordingly undertook an evidence-based, multidisciplinary, iterative process that resulted in an electronic PtDA for older adults with NSTEMI. To our knowledge, *NSTEMI Decide* is the first PtDA developed for this condition in older adults.

When evaluating criteria for effective PtDAs, *NSTEMI Decide* meets all of the key development components as outlined in the IPDAS criteria,^44^ including an iterative development process, presenting available treatment options, providing probabilities of outcomes supported by the most up to date evidence, addressing emotional and social effects, and delivering the PtDA in plain language. We note that some IPDAS effectiveness components (e.g. field testing with users and its relevant subcomponents) are not yet met given the current stage of our development process.

Through a systematic and iterative development process with input from patient and clinician advisory groups, we incorporated a user-centered design that considered multiple perspectives. Preferences for language choice and PtDA design sometimes differed: for example, while clinicians preferred to present a range of risk estimates to ensure the data were inclusive of the older adult target population, patients preferred a definitive and “straight to the point” presentation of information. Additionally, while clinicians preferred to provide study information to justify risk estimates, patients found it overwhelming to grapple with the nuances and limitations of individual research studies. This deliberation between less versus more detail content in PtDAs has been previously documented by others.^25^ Accordingly, we worked to ensure a balance between simplicity and accuracy by utilizing an approach of graphical risk presentation that is accompanied by short, easy-to-read explanatory narratives.

In comparison with PtDAs for other cardiovascular conditions, our PtDA is unique in its attention to design for older adults, where age-related cognitive, visual, and hearing challenges are common. This is especially important given that nearly 30% of U.S. older adults are classified as being below basic health literacy.^45^ As also suggested by our advisory groups, we therefore placed considerable effort on incorporating simple illustrations, recognizable icons, high contrast colors (that were also interpretable by people who were colorblind), and large font into our PtDA to increase readability and knowledge.

There are several limitations to our study that warrant consideration. First, patient advisory groups were from the NYULH referral network, and therefore may not represent regional or social differences in culture and medical decision making. Additionally, while we sought opinions from a variety of clinicians on our risk estimates (invasive and noninvasive cardiologists, general internists, geriatricians), estimates of absolute risk and benefit from invasive coronary angiography are likely to vary widely between clinicians, and are further modified by factors such as very advanced age and comorbidity burden. Further, we limited evidence of benefit described in the PtDA to data from randomized controlled trials, which are rare in this population; while the ongoing SENIOR-RITA trial will provide further definitive data^46^, it is not expected to be completed for several years. In addition, our estimates of risk were not individualized given the paucity of data to make this possible. Lastly, *NSTEMI Decide* still needs to be tested in a prospective study to understand broader usability and impact. In this context, we are currently piloting our PtDA in the inpatient setting to evaluate feasibility of use, satisfaction with care, medical knowledge, decisional conflict, and trust in the medical team.

## CONCLUSION

In conclusion, to our knowledge we developed the first PtDA to be used for older adults hospitalized with NSTEMI. This tool incorporates perspectives from both patients and clinicians, and was largely designed based on consensus (IPDAS) criteria for PtDA development. While prospective validation will be necessary, we believe this PtDA can play an important role in helping care for the growing population of older adults with NSTEMI facing the decision of whether to undergo invasive coronary angiography.

## Data Availability

The authors confirm that the data supporting the findings of this study are available within the supplementary materials. Additional data that support the findings of this study are available on request from the corresponding author, JAD.

## ACKNOWLEDGMENTS

We thank Lysy Gonzalez and Andrea Peña for translating NSTEMI Decide into the Spanish language, and Anna Kiefer for programming the decision aid into a mobile application.

## Author Contributions

Study concept and design: Summapund, Sibley, Kiefer, Iqbal, Spatz, Barnett, Chaudhry, Dickson, Matlock, Dodson.

Acquisition of subjects/data: Summapund, Sibley, Kiefer, Rivers, Langford, Sikand, Dodson. Analysis and interpretation of data: Summapund, Sibley, Iqbal, Spatz, Chaudhry, Dickson, Sikand, Matlock, Dodson.

Preparation of manuscript: Summapund, Sibley, Sikand, Dodson.

## FIGURE LEGENDS

**Supplemental Table S1**. Evidence of Available Treatment Options for Older Adults Hospitalized with NSTEMI

