## Supplemental Table S1 for "NSTEMI Decide: Development of a Decision Aid for Older Adults with Non ST Elevation Myocardial Infarction"

**SUPPLEMENTAL MATERIAL**

**Supplementary Table S1. Evidence of Available Treatment Options for Older Adults Hospitalized with NSTEMI**

**Randomized controlled and meta-analysis data with relative benefit and harm**

| Study | First Author | Year | Total Patients | Duration (Months) | Population | Design | Relative Benefit | Relative Harm |
| --- | --- | --- | --- | --- | --- | --- | --- | --- |
| FIR (FRISC, ICTUS, RITA-2) | Damman | 2012 | 839 | 41.6 | ≥ 75 yo presenting with NSTEMI or UA | Collaborative analysis of individual data from FRISC, ICTUS, RITA-2 RCTs. Routine invasive + selective invasive PCI for refractory angina or HF) | Significant reduction in MACE (18.3% intervention vs. 15.9% control). HR 0.60, 95% CI 0.43-0.83, p=.002 | No significant difference in major bleeding (2.9% intervention vs. 1.9% control, p=.069). |
| Italian Elderly ACS Study | De Carlo | 2012 | 313 | 12 | ≥ 75 yo presenting with NSTEMI or UA | Early Invasive (angiography with revasc within 72 hours) + initially conservative (angiography with revasc for ischemia only) | Non-significant reduction in MACE (27.9% intervention vs. 34.6% control). HR 0.80, 95% CI 0.52 - 1.19, p=0.2 | No significant difference in major bleeding |
| After Eighty Study | Tegn | 2016 | 457 | 36 | ≥ 80 yo presenting with NSTEMI or UA | Invasive + medical therapy | Significant reduction in MACE (40.6% intervention vs. 61.4% control). HR 0.48, 95% CI 0.37-0.63, p=.0001. NNT 4.8. | No significant difference in stroke or major bleeding (HR 0.61, 95% CI 0.22 -1.60, p=.26). |
| Early Invasive versus selectively invasive strategy in patients with non-ST segment elevation acute coronary | Angeli | 2013 | 9,400 | Variable | ≥ 65 yo | Meta-analysis of 9 RCTs comparing Early Invasive + Selective invasive | Significant reduction in MACE (OR 0.85, 95% CI 0.66-0.87). |  |

**Non-randomized data with relative benefit and harm**

| Study | First Author | Year | Subjects | Follow Up  (Months) | Population | Design | Relative Benefit | Relative Harm |
| --- | --- | --- | --- | --- | --- | --- | --- | --- |
| Effect of an invasive strategy on in-hospital outcome in elderly patients with non-ST segment myocardial infarction | Bauer | 2007 | 1936 | 12 | ≥ 75 yo with NSTEMI or UA | Retrospective analysis angiography vs. conservative treatment | Significant reduction in MACE (OR 0.56, 95% CI 0.38 -0.81, p<.0001) |  |
| Management and 6-month outcomes in elderly and very elderly patients with high-risk-non-ST elevation acute coronary syndromes: The Global Registry of Acute Coronary Events | Devlin | 2008 | 8,086 | 6 | ≥ 70 yo with NSTEMI or UA | Observational data from the GRACE registry: PCI + OMT vs. medical therapy alone | Significant reduction in MACE in elderly >70 yo (OR 0.60, 0.47-0.76); very elderly >80 yo (OR 0.72, 0.54-0.95) | No significant increase in stroke risk |
| Influence of Age on Use of Cardiac Catheterization and Associated Outcomes in Patients with Non-ST- Elevation Acute Coronary Syndromes (ACS I, ACS II, GRACE) | Bagnall | 2009 | 11,742 | 12 | ≥ 65 yo with NSTEMI or UA | Retrospective from ACS I, ACS II and GRACE registries. Early invasive strategy vs. conservative management | Significant reduction in MACE (OR 0.45, 95% CI 0.37-0.56, p<0.001). | No significant difference in major bleeding (p=.13) |
| Improving in-hospital mortality in elderly patients after acute coronary syndrome - A nationwide analysis of 97,220 patients in Taiwan during 2004-2008. | Hseih | 2012 | 47,852 | Variable | > 65 yo with NSTEMI or UA | Retrospective registry data PCI vs. medical therapy | Significant reduction in MACE (OR 0.3 age 75-79, 0.4 age 80-84) |  |
| Quality of life after percutaneous coronary intervention in the elderly with acute coronary syndrome | Li | 2012 | 624 | 12 | > 60 yo with NSTEMI or UA | Prospective, non-randomized PCI vs. medical therapy | Significant improvement in QOL scores (OR 1.79, 95% CI 1.10-2.92) |  |
| The impact of increased age on outcome from a strategy of early invasive management and revascularisation in patients with acute coronary syndromes: retrospective analysis from the ACACIA registry | Malkin | 2012 | 683 | 12 | ≥ 70 yo with ACS | Retrospective early revascularization vs. medical therapy | Significant reduction in MACE (OR 0.4, 95% CI 0.2-0.7) | Significant increase in bleeding (HR 14.9, CI 5.6-40, p=0.01) |
| Early Invasive Versus Initial Conservative Treatment Strategies in Octogenarians with UA / NSTEMI | Kolte | 2013 | 986,542 | Variable | > 80 yo with NSTEMI or UA | Retrospective Early Invasive vs. Medical therapy | Significant reduction of in-hospital mortality (OR 0.76) | Significant reduction in stroke (OR 0.63, 95% CI 0.6-0.66) |
| Sex-Related Outcomes in Elderly Patients Presenting with Non-ST Segment Acute Coronary Syndrome: Insights from the Italian Elderly ACS Study | De Carlo | 2015 | 645 | 12 | ≥ 75 yo with NSTEMI or UA | Retrospective registry and analysis of previous RCT data: Early invasive vs. initial conservative | Significant reduction in MACE (27.6% vs. 38.7%, p<0.01) |  |
| Acute coronary syndrome in octogenarians: association between percutaneous coronary intervention and long-term mortality | Barywani | 2015 | 353 | 41.6 | ≥ 80 yo with ACS | Retrospective PCI vs. medical therapy | Significant reduction in all-cause mortality (54.9% vs. 83.1%, p=.001) |  |
| Invasive versus non-invasive management of older patients with non-ST elevation myocardial infarction (SENIOR-NSTEMI): a cohort study based on routine clinical data | Kaura | 2020 | 1976 | 3 | ≥ 80 yo with NSTEMI | Retrospective propensity score-matched cohort: invasive vs. non-invasive management | Significant reduction in adjusted mortality (HR 0.68, 95% CI 0.55-0.84), significant reduction in heart failure hospitalization (HR 0.67, 95% CI 0.48-0.93) | None reported |
